# A novel method to assess motor planning deficits in patients with Parkinson’s disease and mild cognitive impairment

**DOI:** 10.1101/2024.11.19.24317500

**Authors:** Natalie J Maffitt, Sattwika Banerjee, Subhajit Sarkar, Suchismita Majumdar, Supriyo Choudhury, Hrishikesh Kumar, Demetris S Soteropoulos, Alexander Kraskov

## Abstract

It is well established that patients with Parkinson’s disease (PD) show deficits with movement execution, however experiences of motor planning dysfunction, and how they relate to the severity of motor symptoms, remains unclear. To investigate motor planning in PD, we designed a novel precision-grip task. PD patients showed significantly higher uncertainty in task performance compared to healthy controls, indicative of motor planning deficits. Performance of PD patients did not correlate with indicators of disease severity or subtype, yet patients on a higher daily levodopa dosage showed reduced motor planning deficits. Interestingly, these deficits were present even in recently diagnosed patients, implying that this measure may have potential as an early marker of motor planning impairment. These results suggest that the motor planning deficits revealed by our task may arise from separate pathological processes to that of motor execution dysfunction in PD, though might be alleviated with higher treatment dosages.

## Introduction

Whilst the defining characteristics of people with motor disorders are difficulties in generating and controlling movement, additional features of cognitive dysfunction are also commonly experienced. For those with Parkinson’s disease (PD), this can range from minimal cognitive impairment in, for example, mood regulation or executive function, to extreme clinical pictures of severe dementia ^[1]^. Of these deficits, impairments in motor planning can have consequences for successful interaction with our environment in how they perform day to day physical tasks, yet such impairments have comparatively received much less focus.

Our understanding of motor planning deficits in PD initially developed within the context of action imitation tasks. Sharpe, Cermak and Sax (1983), showed that when asked to perform either familiar gestures from memory or unfamiliar gestures post observation, people with PD only make greater spatial errors compared to controls when performing unfamiliar gestures ^[2]^. It has been suggested that such errors are a consequence of defective encoding and processing of visuospatial information, since the neural mechanisms needed to perform successful action imitation rely more heavily on the accuracy of visuospatial encoding than when an individual is responding to verbal command ^[3,4]^. Yet, these spatial errors could also be influenced by memory recall deficits; individuals with PD show greater impairment when imitating movement sequences compared to single actions ^[3]^. Thus, the motor planning performance of individuals with PD when intending to enact their own self-chosen movement, rather than one rehearsed from memory or by way of imitation (i.e. less reliant on successful visuospatial encoding or recall), remains to be explored.

Additional evidence for the presence of motor planning abnormalities in PD has been implied from reaction time (RT) tasks. For simple RT tasks, whereby the subject knows in advance the target location or required action but is instructed to wait for a go signal before initiating movement, consistent slowing of RT is observed in comparison to healthy controls (e.g. ^[5,6]^. There have been suggestions that this delay may not solely be a manifestation of bradykinesia, but rather could reflect problems in releasing motor commands or in the pre-programming of them ^[7,8]^. Contrastingly, in choice RT tasks, several actions are presented and the one required is only revealed at the time of the go cue. Not only, however, do reported deficits in choice RT tasks remain controversial due to the seemingly task-dependency nature of the findings, but visual processing time, which has been shown to be hindered in PD, is consistently an uncontrolled factor in both forms of RT tasks ^[9-12]^.

In this study, we used the context of motor ambiguity in a reach-to-grasp task as a method to investigate motor planning capabilities in people with PD, in a task adapted from Stelmach et al., 1994 ^[13]^. In this context, motor ambiguity can be described as situations whereby the motor action (i.e. wrist posture) performed upon a target object is variable, despite the visual-spatial properties of the object remaining the same. For a healthy person, the decided motor action for a task, such as picking up an object with the wrist pronated, is decided by individualised comfort preferences, leading to an optimised and consistent action choice based on previous learned experience. However, some motor ambiguity will nevertheless persist when the optimal preference cannot be identified and therefore planned. We hypothesised that when performing a reach-to-grasp task, people with PD would show greater ambiguity in wrist posture selection as a consequence of impaired motor planning in comparison to healthy controls. Given that the characteristics of PD consist of both motor and non-motor deficits, it was of interest to also investigate motor planning performance in individuals who experience cognitive dysfunction without motor execution deficits. We therefore recruited subjects diagnosed with mild cognitive impairment (MCI), a cohort that experience cognitive deficits in memory, language and other cognitive domains, though without significant daily disruption to be classified as dementia and notably absent of any basic motor deficits ^[14]^. It has been shown previously that individuals with MCI may also experience visual-motor difficulties when their cognitive load is high: they are slower to plan movements and have difficulties integrating visual perception with motor action, though in simpler cognitive tasks such as standard reach-to-grasp, performance and planning deficits are less pronounced ^[15-17]^. In this study, we explored the possibility that in pre-dementia, the extent of motor ambiguity may reflect the weight of cognitive load induced by our grasping task and thus reveal any impairments in motor planning; we hypothesised any such deficits will occur to a minimal extent given the predictive cognitive simplicity of the task.

We reveal not only that motor planning is affected in both PD and MCI in comparison to healthy controls, but also that for those with PD it can be influenced by daily treatment dosage with levodopa, suggestive of a therapeutic influence of dopamine on motor planning.

## Methods

### Participants

This study was conducted in a total of 60 participants within the Neurology Outpatient Department of a tertiary care referral centre in Eastern India. Ethical approval was obtained from the Institutional Research Ethics Committee of the Institute of Neurosciences, Kolkata; written informed consent was taken from all volunteers. This study was performed in accordance with the guidelines established in the Declaration of Helsinki, except that the study was not preregistered in a database. Healthy volunteers were selected from the care givers of the patients attending the Neurology Outpatient Department; all had normal cognition (MOCA ≥ 26), and no apparent neurological disease. People with PD were diagnosed using UK Brain Bank diagnostic criteria for PD; exclusion criteria for participation in this study included presence of dyskinesia, severe cognitive impairment (MOCA < 10), and profound physical impairment (Hoehn & Yahr stage > 4) or an inability to perform prehension movements. Inclusion criteria for the MCI cohort consisted of a MOCA score indicating mild cognitive impairment at the time of assessment (MOCA < 26 and > 10) and a complete absence of any signs of motor execution dysfunction. All participants were right-handed and naïve to the experiment and its predictions.

Due to low trial counts (< 50% complete), 7 participants (6 PD, 1 Control) were excluded from analysis. Remaining in the study were 15 healthy controls (8 females; 51.8 ± 9.6 years STD), 27 people diagnosed with idiopathic PD (IPD; 12 females; 60.9 ± 10.7 years), and 11 diagnosed with MCI (5 females, 63.5 ± 7.5 years). All PD participants were receiving regular levodopa treatment (mean daily levodopa equivalent dosage (LED) 511.0 ± 278.6 mg) and assessed in their ON state. Conversion formulae for LED calculation were in accordance with widely accepted proposals ^[18]^.

### Clinical scoring

The disease severity of each participant was assessed using standard disease severity scales: for MCI, the Montreal Cognitive Assessment (MOCA); for PD, the Movement Disorders Society Unified Parkinson’s Disease Rating Scale-III (MDS-UPDRS III) and MOCA. The MOCA and/or MDS-UPDRS assessment were performed immediately preceding or following the main task by a trained professional. The MOCA scores were corrected for education level.

### Task set-up

The experimental paradigm required participants to perform a precision grip movement on a triangular prism block (6 cm long by 2 cm wide, see Figure 1 and previous paper ^[19]^) with their right-hand upon hearing a go cue (auditory beep 500 Hz, lasting 50 ms). The target object was designed with the intention to afford only one possible grasp: the thumb and finger at opposing ends of the prism block ^[13]^. The object was mounted onto a grey rectangular stage and connected to a stepper motor to enable rotation in steps of 0.9° angles - the stepper motor was driven by an Arduino UNO R3 and motor shield R3, and controlled by custom written Arduino scripts. Thin triangular metal plates were glued to each end of the prism block and connected to a custom made circuit that allowed detection of skin contact by a change in impedance. By placing a glove onto the index finger to impede contact with the object, hand orientation selection could be recorded in terms of thumb placement, either ‘Grasp 1’ whereby the thumb was placed on the left or top end of the object or ‘Grasp 2’ whereby the index finger was used instead (Figure 1). Previously studies have referred to these two grasp postures as ‘thumb-left’/’supination’ or ‘thumb-right’/’pronation’ respectively (e.g. Stelmach et al., 1994; Wood and Goodale, 2011), however given both of these nomenclatures lose validity at certain positions we opted for Grasp 1/Grasp 2.

**Figure 1.**
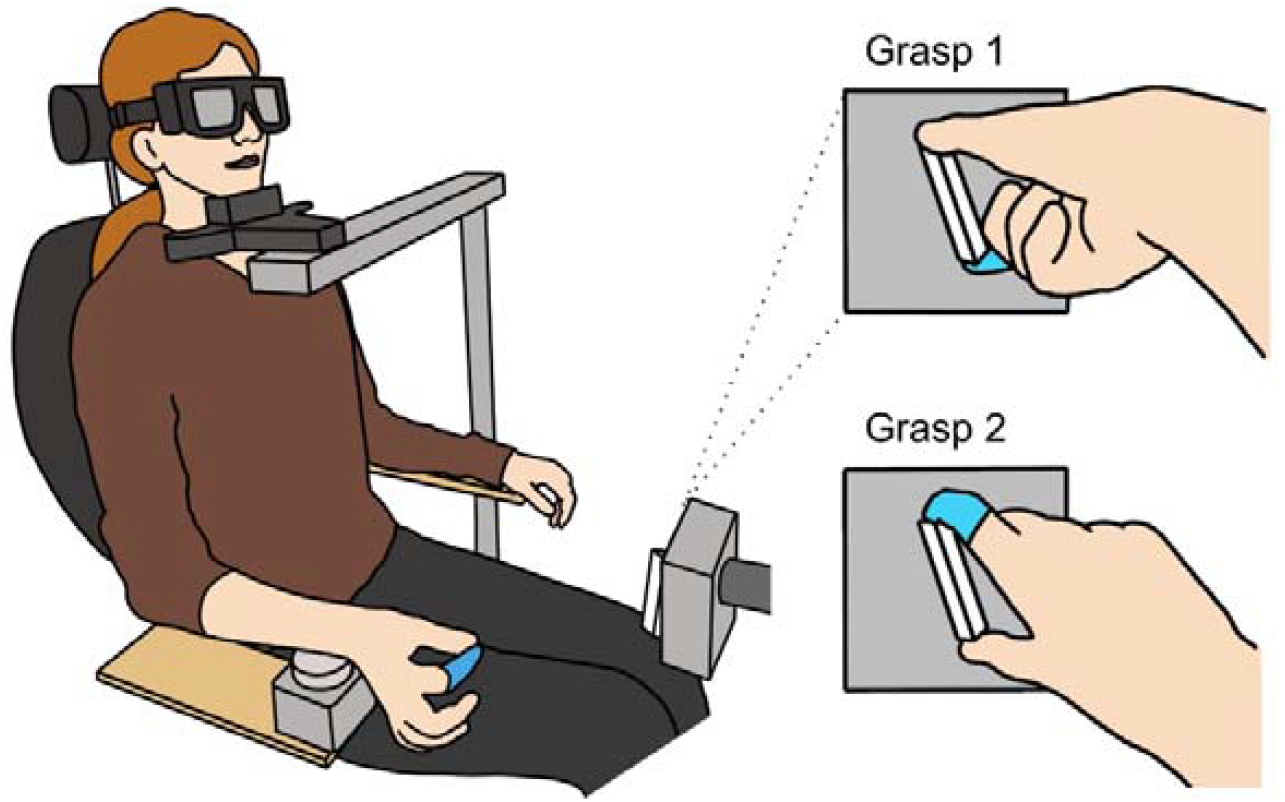
Experimental set-up. Participant is seated with LCD goggles covering their visual field and head placed on a chin rest. Their right wrist is used to press the home-pad button to initiate trial sequence. A latex glove on the index finger impedes touch with the contact circuit on the target object, allowing detection of wrist posture (Grasp 1 or Grasp 2) on a trial-by-trial basis.

Participants were seated in a chair with their wrist positioned on a circular ‘home-pad’ button (6 cm in diameter) as the starting location. The target object was positioned 31 cm anterior to the centre of the home pad and 14.5 cm superior, with its rotation occurring in the frontal plane. This resulted in the object lying 27 cm to the right of the body’s midline.

As comfort and therefore choice of grasp has been shown to be sensitive to body and head posture ^[20]^, a chin rest was used to mitigate any postural shifting that could introduce unwanted variability to behaviour. Participants wore LCD goggles (PLATO goggles; Translucent Technologies, Toronto, Canada) to occlude their vision between trials until the object had rotated into place at the target angle. The transition time from opaque to visible was 4 ms. In cases where participants wore visual correction glasses, the goggles were placed over the top and secured with a headband.

### Task procedure

Beginning each trial with their wrist in mid-pronation to press down on the home-pad, participants were asked to lightly pinch their index finger and thumb together until the go cue. This wait time was uniformly varied randomly from trial to trial (1000-1300 ms) to prevent participants acting on anticipation of the go cue. Synchronous with an auditory beep, the goggles became transparent and participants were asked to reach out as quick as possible and comfortably grasp the target object between their thumb and index finger, pinching it lightly. After object contact, they returned to the home-pad to initiate the next trial sequence. If the home-pad was released too early (i.e. prior to the go cue), an error tone would sound (200 ms, 200 Hz) to indicate the subject should return to the start button. A total of 162 trials across 27 unique angles spaced by 4.5° (spanning 127.8°) were carried out in a randomised sequence with equal incidence per angle.

### Data and statistical analysis

Grasp choice data were gathered using the custom designed contact circuit and sampled at 5 kHz (CED Micro 1401, Cambridge Electronic Design) using Spike2 software. All data analyses and statistics were performed using custom written MATLAB scripts. Analysis resulted in three measures – the halfwidth (as described below), reaction time (RT), measured as time from go cue to home pad release, and movement time (MT), measured as time from release of home pad to contact with the target object. Trials for which RT was shorter than 150 ms were excluded from analysis; MTs had no explicit cut-off. For both RT and MT analysis, remaining values were log transformed, with any trials outside the mean ± 3 SD excluded - this exclusion process was then repeated recursively until no more trials could be excluded. On average across all participants, 3.3 % of the trials were discarded due to extremes of RT and 0.91 % for extremes in MT.

Choice of grasp was coded with reference to which edge of the target object the thumb made contact with. The probability of using a ‘Grasp 2’ wrist posture was calculated for each target angle and a logistic regression curve was fitted to the data using the function below, where *x*_o_ is the subject’s switch point between the two choices, and k is a scaling parameter.

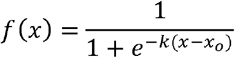

This sigmoid fitting allowed the ‘halfwidth’ to be calculated - a measure of switch sharpness, calculated as the difference between target angles for the 75% and 25% choice probabilities (= *2ln(3)/k*). This was used as an indicator of subject uncertainty across the task as a whole.

Across each group population, numerical values were described using mean (± SD) and compared using one-way ANOVA with post hoc Tukey’s test. The strength of correlation was estimated with Pearson’s correlation coefficient R^2^. For all three variables (HW, RT, MT), receiver operating characteristic (ROC) curves were constructed and the area under the curve (AUC) calculated. An AUC of 0.5 indicates an identical distribution of the measure in patients and controls; an AUC of 1.0 occurs if the distributions are completely non-overlapping. Thus, a higher AUC value indicates greater utility of the measure as a diagnostic tool. Comparison of AUC values was performed using DeLong’s test.

## Results

Figure 2 shows the average decisions in grasp choice for each target angle from single representative subjects of each cohort, which illustrate the first main finding of our study. As previously reported, when the target object is rotated across a range of angles, a typical healthy subject transitioned from consistently using one grasp posture to using the other across a relatively narrow range, following a sigmoid pattern (Figure 2a). The steepness to the sigmoid slope in this healthy subject is a consequence of minimal variability in grasp choice across most angles, yielding a halfwidth value (indicated by arrows) of 12.1° . By contrast, for the individual with IPD, there are greater target angles for which grasp choice is not consistent, leading to a wide ambiguity zone with a halfwidth value of 34.6° (Figure 2b). Similarly for the single subject with MCI (Figure 2c), variability in grasp choice occurs over a wider range of angles, yielding a larger halfwidth value (19.3°) than the example healthy control.

**Figure 2.**
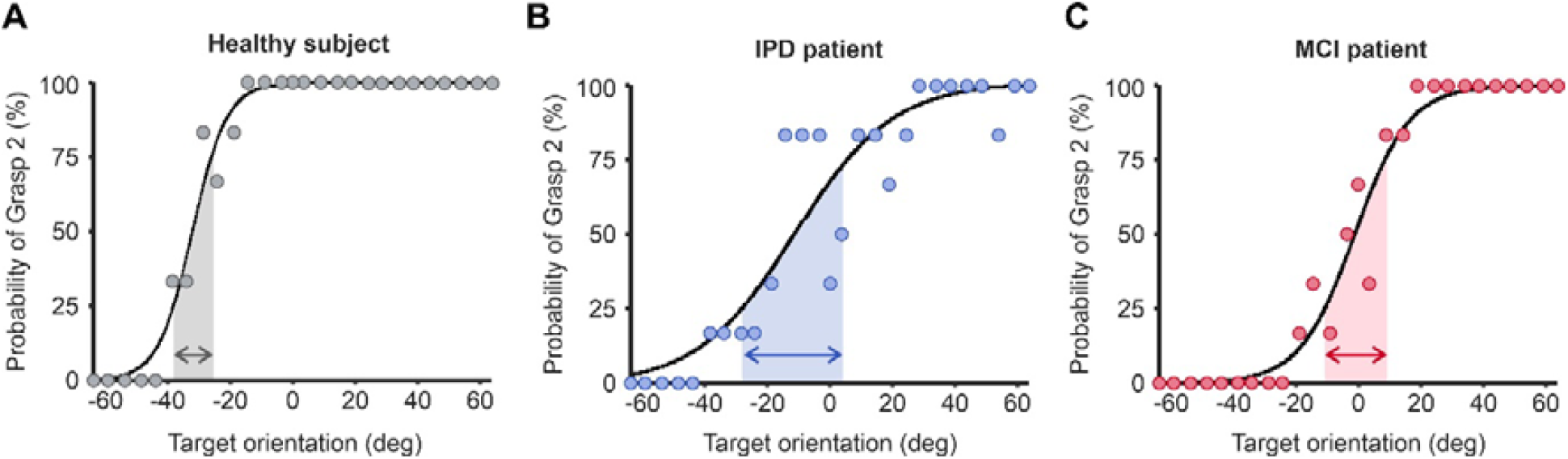
Example psychophysics curves from an individual from each cohort. Shaded areas indicate the range of angles for which the probability of using a Grasp 2 posture over Grasp 1 is between 25 and 75%, yielding the measure ‘Halfwidth’. IPD, idiopathic Parkinson’s disease; MCI, mild cognitive impairment.

For all subjects within each group, the halfwidth of the sigmoid curve was calculated, shown by each individual dot in Figure 3a. The healthy control cohort had a lower mean halfwidth value than each patient group (Figure 3a; Control mean = 12.86 ± 5.09 °; IPD mean = 25.90 ± 8.84 °; MCI mean = 21.76 ± 7.04 °; ANOVA p < 0.0001). Post-hoc t-tests between the healthy control group and both IPD and MCI cohorts indicated significant differences in halfwidth (IPD p < 0.0001; MCI p = 0.049), however there was no significance difference between the IPD and MCI cohorts (p = 0.098).

**Figure 3.**
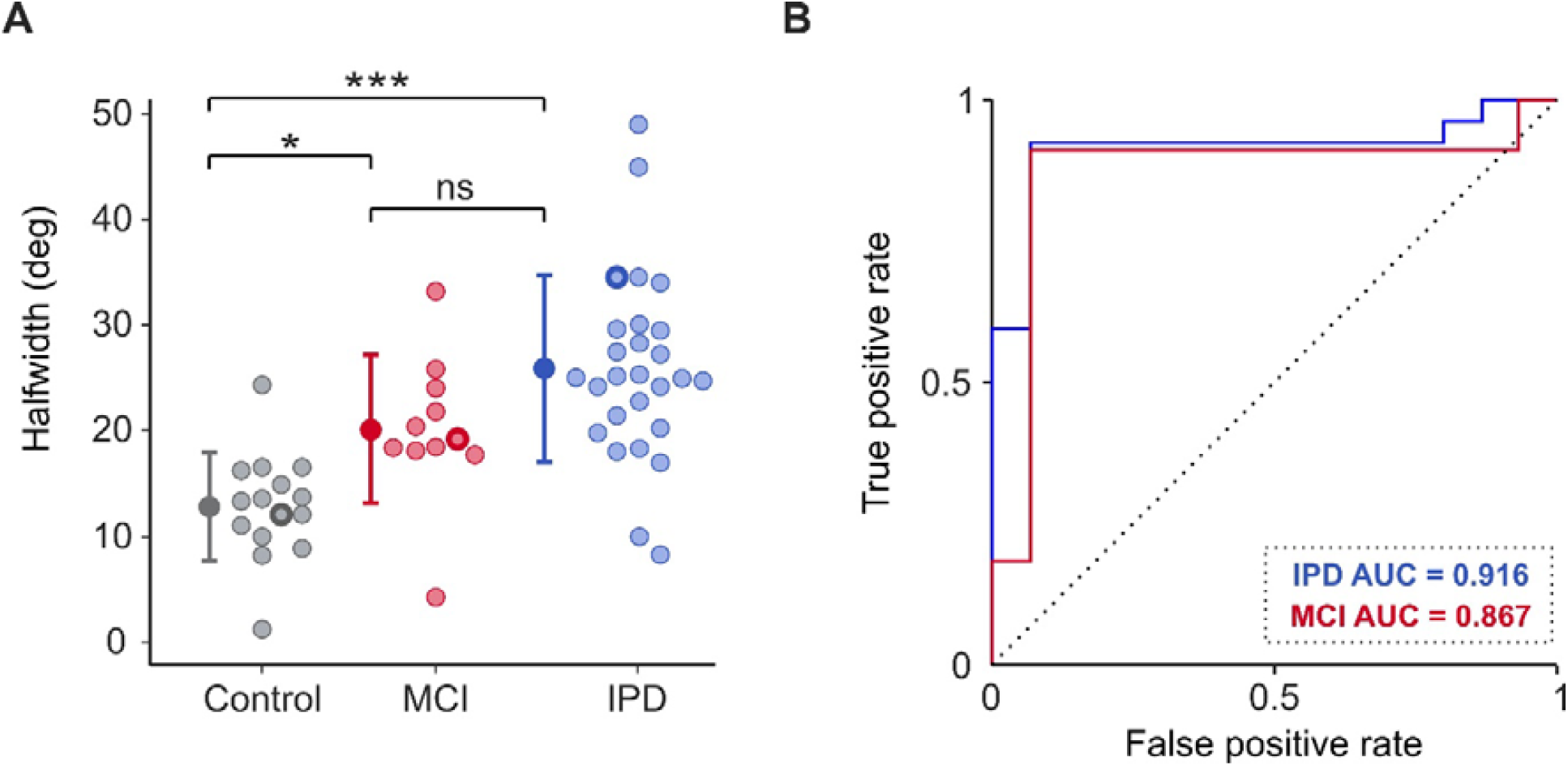
Halfwidth values and receiver operating characteristic (ROC) curves for control, PD, and MCI subjects. a) Halfwidth values for every participant in each cohort. The example data shown in Figure 2 is highlighted here by thicker outline on the individuals’ data point. b) ROC curves confirming that halfwidth can effectively separate healthy controls from both PD and MCI individuals. Dotted diagonal line indicates the expected result if halfwidth did not discriminate between groups. IPD, idiopathic Parkinson’s disease; MCI, mild cognitive impairment. Error bars represent mean ± 1 STD. * P < 0.05; *** P < 0.0001; ns, not significant.

Given halfwidth was significantly different between each patient group and healthy controls, it was of interest to investigate whether this difference was significantly robust to distinguish motor planning pathology from health. This is explored in Figure 3b, which presents receiver operating characteristic (ROC) curves for both IPD and MCI cohorts. The area under the ROC curve for HW was 0.916 and 0.867 for IPD and MCI cohorts respectively; both values were significantly higher than 0.5, the value associated with entirely overlapping distributions, (p < 0.005 for both, Monte Carlo test, 10,000 iterations).

To explore whether the observed difference in PD from controls was related to disease severity or clinical subtype, MDS-UPDRS assessment was carried out. Interestingly, there was no significant correlation between MDS-UPDRS part III score and sigmoid halfwidth (Figure 4a; R^2^ = 0.014, p = 0.551). Severity of bradykinesia, calculated from a subset of the MDS-UPDRS (Supplementary Data 1), also showed no significant correlation with halfwidth (Figure 4c; R^2^ = 0.054, p = 0.787). Furthermore, there was no significant difference in halfwidth between those of Hoehn and Yahr stage 1, and those of stage 2 (Figure 4b; Stage 1 mean = 25.04 ± 6.09; Stage 2 mean = 29.38 ± 11.93; unpaired t-test p = 0.248). Nor did the clinical subtype of PD (tremor dominant – TD, versus postural and gait instability dominant - PIGD) have any significant bearing on halfwidth (Figure 4d; PIGD mean = 22.37 ± 6.40; TD mean = 27.98 ± 9.57; unpaired t-test p = 0.112).

**Figure 4.**
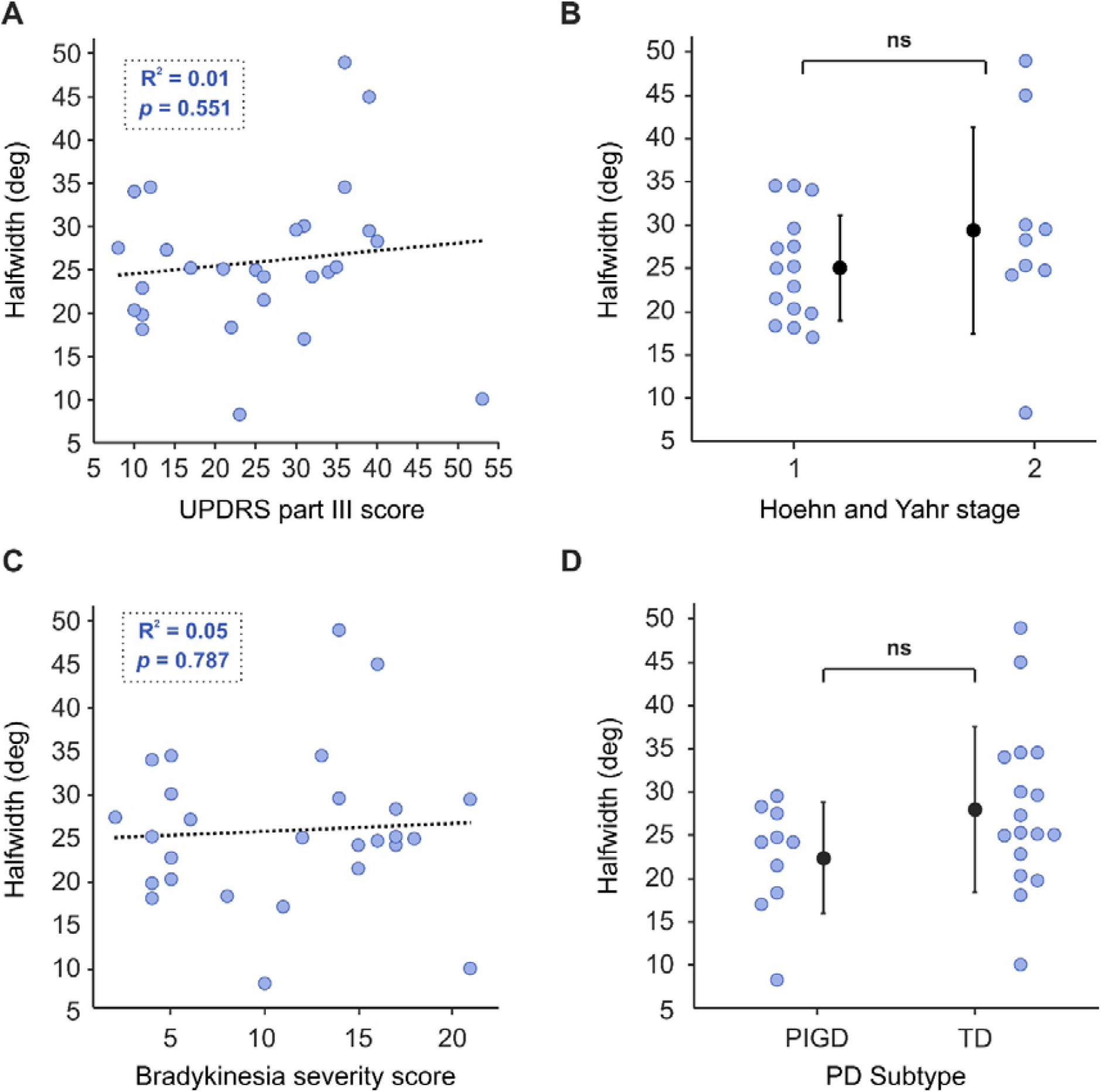
Severity scales and clinical subtype for people with PD. Data is unavailable for four PD individuals for Hoehn and Yahr stage, and eight for time since diagnosis. Clinical subtype is categorised based on MDS-UPDRS scoring. IPD; idiopathic Parkinson’s disease; TD: tremor dominant; PIGD: postural and gait instability dominant; ns: no significance. Error bars represent mean ± 1 STD.

Despite finding no correlation between severity of motor symptoms and halfwidth, surprisingly for individuals with PD there was a significant effect of daily levodopa equivalent dosage (LED) on sigmoid halfwidth (Figure 5a; R^2^ = 0.29, p = 0.004). Interestingly, the higher the daily treatment dosage, the lower the halfwidth value and closer to the mean healthy control value (dashed grey line) subjects became. However, there was no correlation between halfwidth and time since diagnosis (Figure 5b; R^2^ = 0.01, p = 0.634).

**Figure 5.**
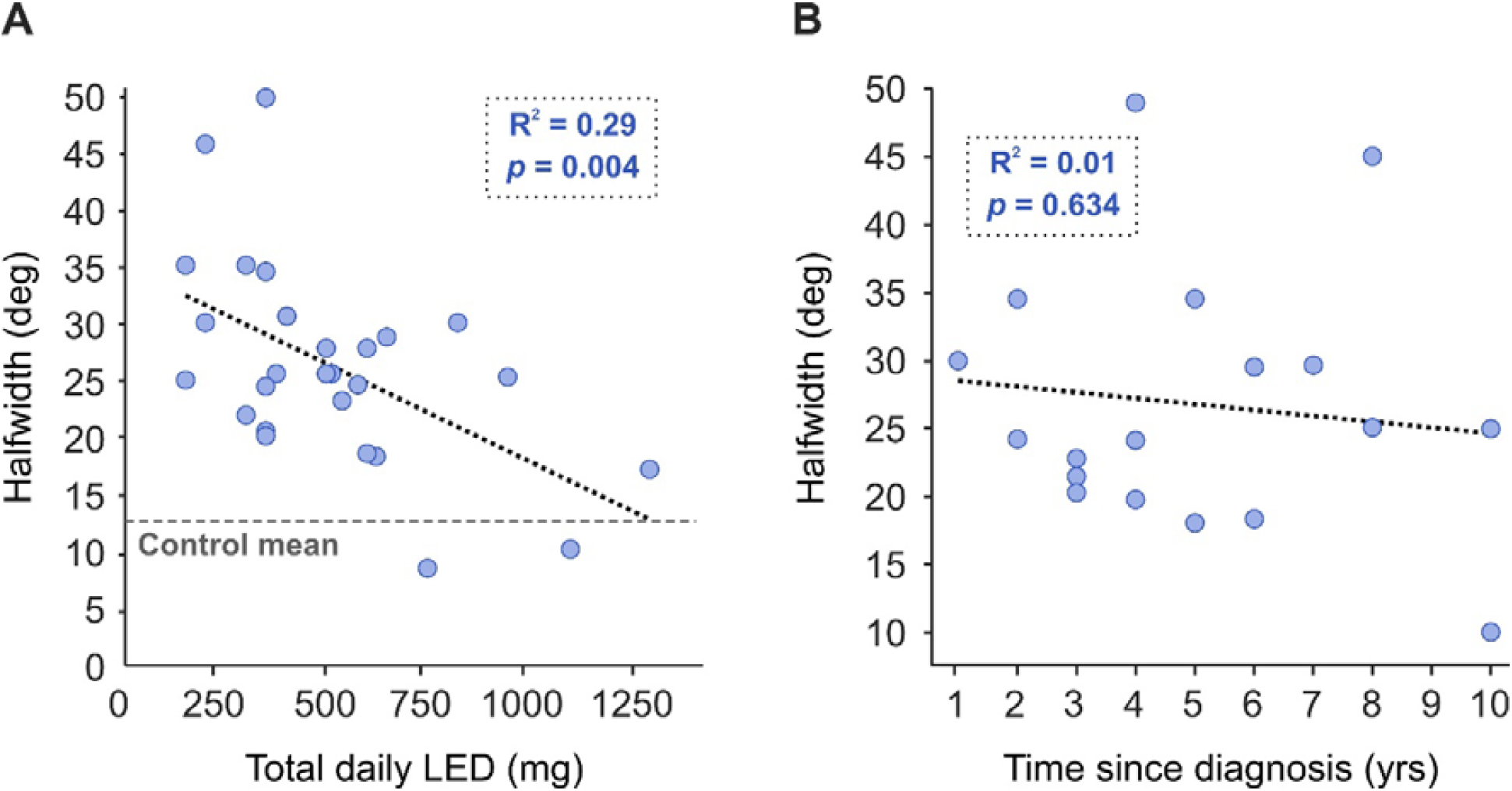
Effect of levodopa and time since diagnosis on halfwidth for people with PD. Total daily levodopa equivalent dosage (LED) was calculated for each subject receiving treatment.

Though LED correlated with our halfwidth measure, it did not correlate with RT (Figure 6a; R^2^ = 0.05, p = 0.250) or MT (Figure 6b; IPD R^2^ = 0.06, p = 0.225).

**Figure 6.**
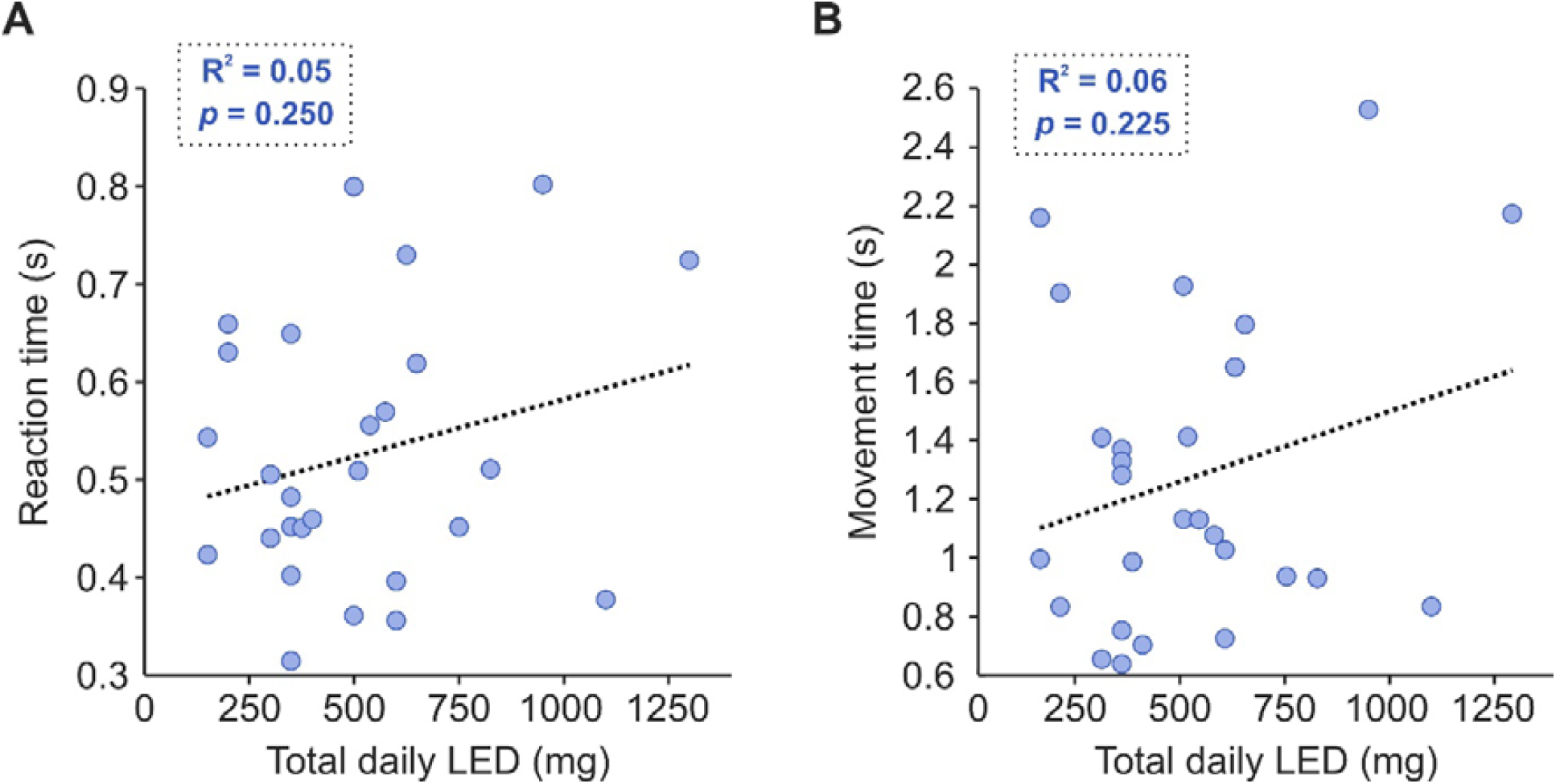
Effect of levodopa on reaction and movement times for people with PD. Total daily levodopa equivalent dosage (LED) was calculated for each subject receiving treatment. RT, reaction time; MT, movement time.

The effect of global cognitive score and age on halfwidth are shown in Figure 7b, 7d, and 7f, and 7a, 7c and 7e, respectively. The age of the subject had no significant influence on their grasp choices for any cohort (Figure 7e, 7c, 7a; IPD R^2^ < 0.01, p = 0.838; MCI R^2^ = 0.01, p = 0.728; Controls R^2^ = 0.01, p = 0.669). Notably, for individuals with MCI, there was a significant correlation between MOCA score and halfwidth (Figure 7d; R^2^ = 0.76, p < 0.001), with a higher MOCA score (i.e. stronger cognitive abilities) increasing the likelihood of a smaller halfwidth value. However, for both the PD cohort and healthy controls this relationship was not found (Figures 7f, 7b; IPD R^2^ < 0.01, p = 0.883; Control R^2^ = 0.05, p = 0.446).

**Figure 7.**
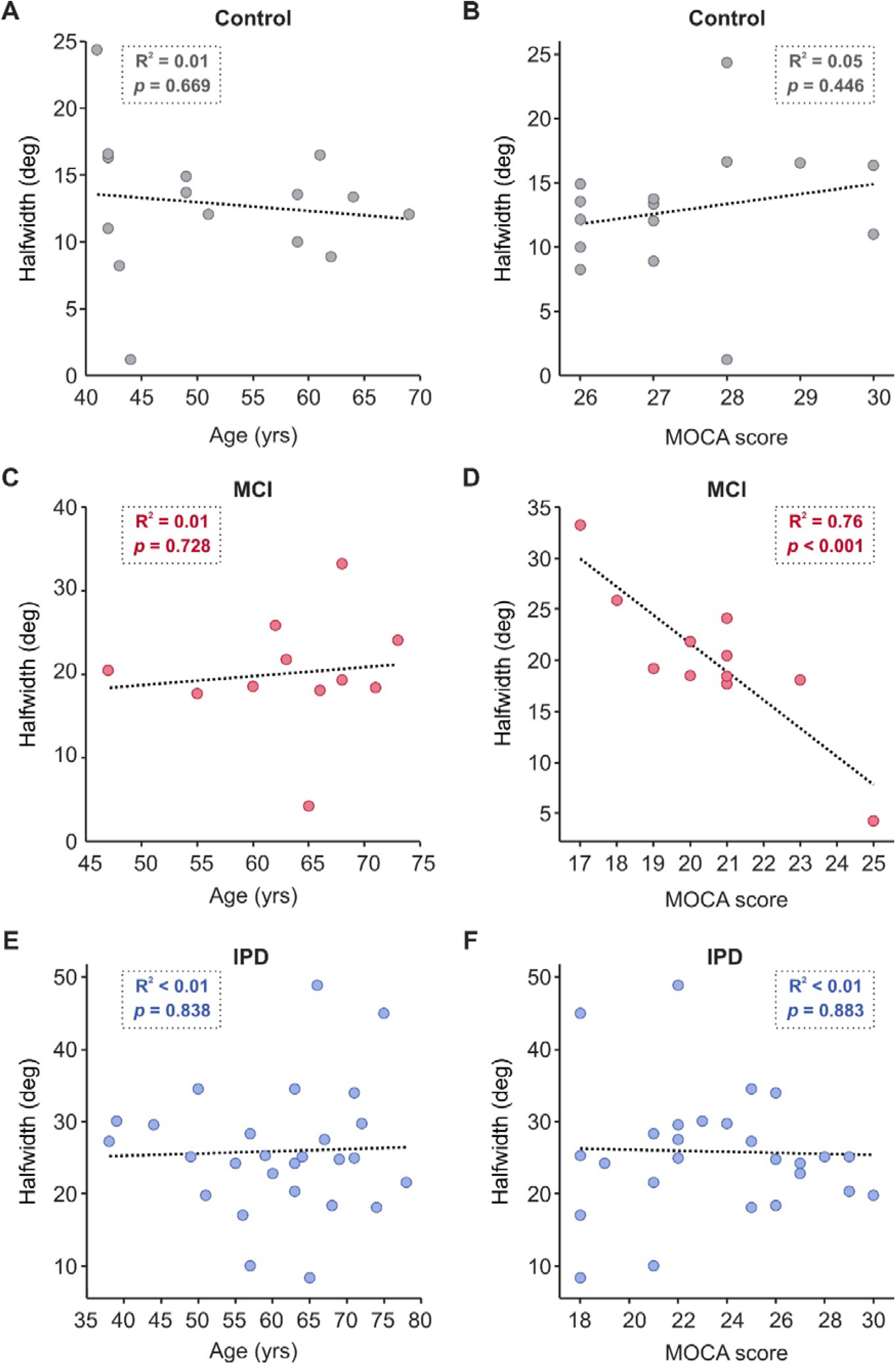
Effect of age and cognition on halfwidth for all cohorts. MOCA subdomain scores for each participant of the IPD and MCI cohorts can be found in Supplementary Data 1. IPD, idiopathic Parkinson’s disease; MCI, mild cognitive impairment; MOCA, Montreal cognitive assessment.

The MCI and IPD cohorts were further described into specific cognitive profiles based on MOCA scoring ^[21]^. Nearly all (10/11) those with MCI showed amnestic traits, scoring ≤ 50% on the memory subdomain (Supplementary Data 1). By the same cut-off criteria, seven showed dysexecutive function, two visuospatial deficits, one attentional deficits, and one orientational deficits. No subject scored ≤ 50% on the language subdomain. For those with IPD, over half displayed dsyexecutive function (14/27), 11 amnestic traits, 5 attentional deficits, and 4 visuospatial deficits. No individual scored ≤ 50% on either language or orientation. Given the complex cognitive profiling for both cohorts and limited sample numbers, it is difficult to disentangle the potential contribution of each subdomain to task performance. However, when pooling scores across both MCI and PD cohorts, no correlation was found between any subdomain and halfwidth scores.

As expected, healthy controls were quicker to release the home-pad upon receiving the go-cue compared to all other clinical disease groups (Figure 8a; healthy mean RT = 0.39 ± 0.05 s; MCI mean RT = 0.51 ± 0.11 s; IPD mean RT = 0.52 ± 0.14 s). An ANOVA indicated a significant effect of group (healthy, MCI, IPD; p = 0.002) on RT, with post-hoc t-tests indicating significant difference between healthy controls and both IPD (p = 0.001) and MCI (p = 0.029) cohorts. No significant difference in RT between IPD and MCI was found (p = 0.883).

**Figure 8.**
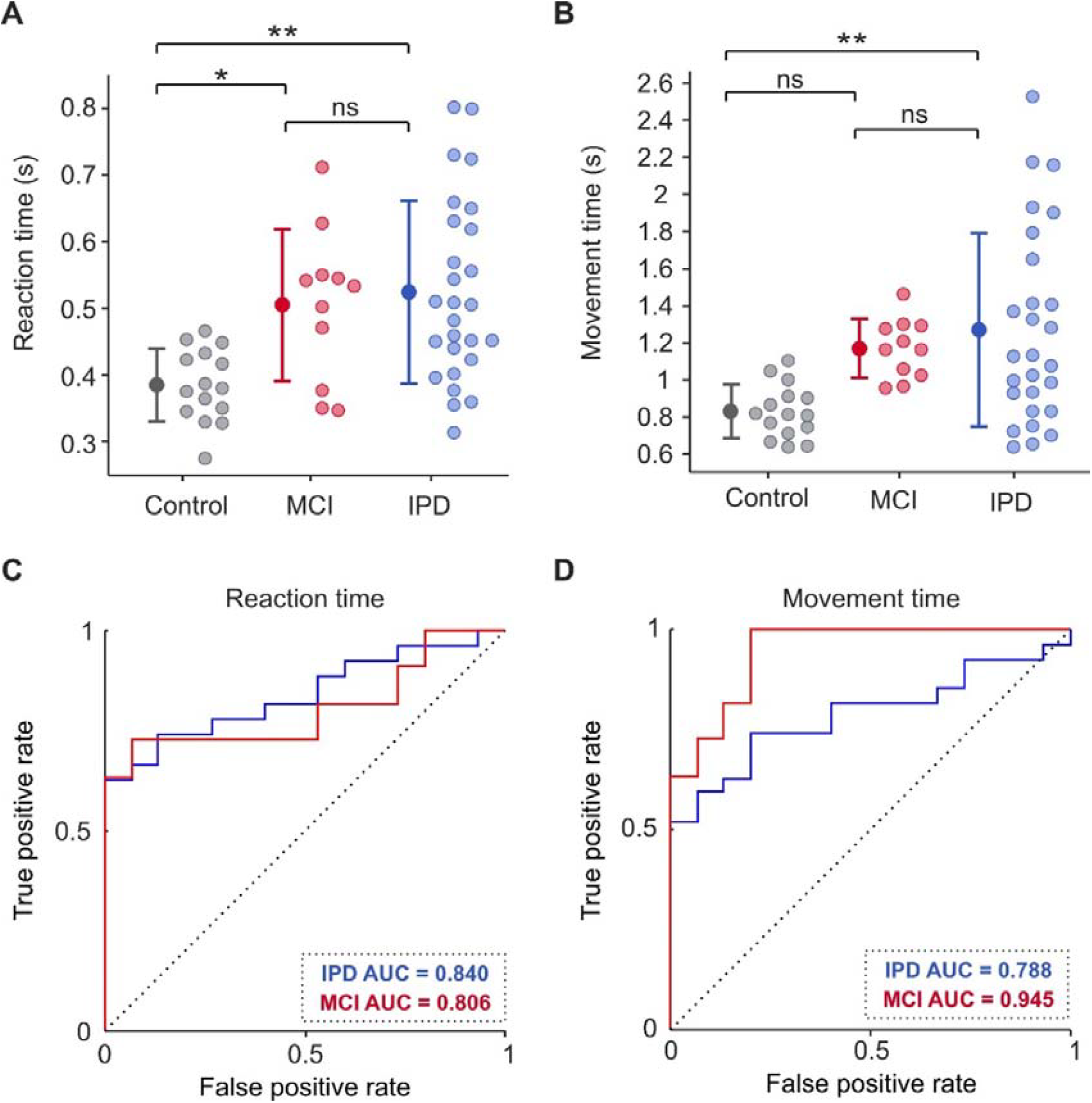
Reaction time (RT) and movement time (MT) values and receiver operating characteristic (ROC) curves for control, PD, and MCI subjects. a-b) RTs and MTs of all control, PD, and MCI subjects. c-d) ROC curves confirming that RT and MT can effectively separate healthy controls from both people with PD and those with MCI. Dotted diagonal line indicates the expected result if halfwidth did not discriminate between groups. IPD, idiopathic Parkinson’s disease; MCI, mild cognitive impairment. Error bars represent mean ± 1 STD. * P < 0.05; ** P < 0.005; ns, not significant.

Healthy subjects were also quicker to move from the home-pad to make contact with the target object compared to both IPD and MCI groups (Figure 8b; healthy mean MT = 0.83 ± 0.14 s; IPD mean MT = 1.27 ± 0.52 s; MCI mean MT = 1.17 ± 0.16 s). An ANOVA indicated a significant effect of group (healthy, MCI, IPD; p = 0.004) on MT, with post-hoc t-tests indicating significant difference between healthy controls and the IPD cohort (IPD p = 0.003), but not MCI (p = 0.082). In addition, no significant difference in MT was found between IPD and MCI (p = 0.759).

To assess the capabilities of RT and MT as diagnostic indicators, ROC curves were compiled, shown in Figure 8c-d. The area under the ROC curve for RT was 0.840 and 0.806, and for MT was 0.788 and 0.945 for IPD and MCI individuals respectively. All values were significantly higher than 0.5, the value associated with entirely overlapping distributions, (p < 0.005 for all, Monte Carlo test, 10,000 iterations). For comparison, there was no significant difference in AUC value between HW and RT either cohort (IPD Z = 1.30, p = 0.193; MCI Z = 0.38, p = 0.702; DeLong’s test), nor between HW and MT (IPD Z = 1.57, p = 0.116; MCI Z = -0.81, p = 0.419; DeLong’s test).

Furthermore, there was no significant correlation between RT and halfwidth for any cohort (Control R^2^ = 0.10, p = 0.261; IPD R^2^ = 0.05, p = 0.278; MCI R^2^ = 0.09, p = 0.365), nor between MT and halfwidth (Control R^2^ = 0.02, p = 0.646; IPD R^2^ = 0.01, p = 0.639; MCI R^2^ = 0.02, p = 0.71). Interestingly, there was also no correlation between RT and bradykinesia severity for those with IPD (R^2^ = 0.15, p = 0.454), nor between MT and bradykinesia severity (R^2^ = 0.25, p = 0.201).

## Discussion

In this study we introduced a novel approach for assessing motor planning capabilities in people with Parkinson’s disease and mild cognitive impairment. Here, the presentation of a target object at varying degrees of ambiguity in a simple reach-to-grasp task allowed us to probe motor planning uncertainty by measuring grasp selection, an output of distinct motor plans. In agreement with previous studies, we observed significant deficits in patients’ task performance relative to healthy controls suggestive of impairments in motor planning, in our case measured by the degree of uncertainty in wrist posture selection. However, unlike previous studies, the context of motor ambiguity in our task allowed the assessment of motor planning capabilities without reliance on action imitation, memory recall, or rapid visual inspection, skills that have been required in previous motor planning assessments ^[2,3,11,12]^. This, combined with evidence that greater uncertainty is present even in those with a more recent diagnosis and with relatively minor cognitive impairment, suggests that grasp uncertainty is a sensitive measure of motor planning impairment that could have utility as an early disease marker.

We found that the degree of uncertainty did not correlate with Hoehn and Yahr stage or MDS-UPDRS (Part III) score, both established scales of PD disease progression scored by the extent of the severity of motor symptoms. It is therefore likely that grasp uncertainty, rather than being a direct consequence of motor execution impairment, actually captures a more cognitive deficit, specifically motor planning. This is further evidenced by the lack of any significant difference in planning uncertainty between tremor dominant, and postural and gait dominant clinical phenotypic subtypes, as well as bradykinesia severity. Cognitive dysfunction is frequently documented in PD individuals at all stages of the disease, yet correlations with objective motor symptom scoring can be highly variable. For example, MDS-UPDRS (Part III) is often found to be a weak or non-existent correlate to both global and specific measures of cognitive impairment ^[22-26]^. In addition, the lack of correlates between cognitive scoring and objective measures of motor control functioning ^[24,27-29]^, and the presence of prodromal non-motor deficits prior to diagnostic motor symptoms ^[30,31]^, all exemplify the occurrence of cognitive deficits and their independence from other pathological changes in PD.

The consequences of disease pathology likely consist of a multitude of cortical and subcortical network changes that lead to further impairment beyond basic motor dysfunction, varying both between individuals and over time. Non-motor symptoms can arise not only independent from the severity of basic motor dysfunction as described above, but also independent from each other ^[32-34]^. While deficits in memory, attention, executive and visuospatial function are common, there exists a large heterogeneity to the profile of cognitive dysfunction in PD ^[35]^. Our finding of the lack of correlation between motor planning deficits and MOCA scores, a measure of global cognitive function ^[21]^, is therefore not unique, but instead builds towards an argument for distinct dysfunctional neural circuitry or neurochemical systems ^[24]^. Depending on what elements of network circuitry become impaired and the nature of interactions between them, different combinations of behavioural deficits could emerge to different extents, hence yielding the absence of correlation between our measure and MOCA or MDS-UPDRS.

Despite the lack of correlation with disease severity scoring, subjects who were receiving a higher total daily treatment dosage showed reduced motor planning deficits compared to those on lower dosages. Given that dosage is determined by the clinician primarily on the basis of the severity of motor symptoms ^[36]^, one could initially interpret LED as a disease severity indicator. However, this would be presumptuous given that dosage can fluctuate based on other clinical considerations such as the severity of negative side effects ^[37]^. Thus, one could consider LED as a proxy for disease management strategy, however this is difficult to interpret without a longitudinal study to explore how management affects motor planning over time. One speculative hypothesis of this relationship between LED and uncertainty could be that lower dosages of levodopa may be insufficient to reduce motor planning deficits, meanwhile higher dosages may be able to restore impaired processing. This potential amelioration may occur to a separate extent than that of improvements to motor function, as implied by the lack of correlation between dosage and MT. That being said, to be able to interpret this relationship between dosage and motor planning beyond our limited speculation, an ON/OFF study across varying dosages should be explored.

Evidence for a cortical influence of levodopa administration has been accumulated based on drug induced behavioural changes in PD since soon after its introduction as a therapeutic drug ^[38]^. Hypomania, impulse control disorder, personality changes, and hallucinations are but a few examples of the common negative side effects of levodopa, however the pathophysiology remains unclear ^[39]^. Though cortical deficits in PD are typically treated with cholinesterase inhibitors acting on acetylcholine ^[30]^, further evidence for levodopa having a cortical effect has manifested in studies showing improved cognitive performance across a range of clinical assessment scales ^[40-42]^, although benefits are not always universal ^[43]^. Nevertheless, these studies have primarily focussed on ON/OFF states rather than comparative dosing effects. As the latter remains unknown, it could be speculated that, based on our findings, some dysfunctional neural networks may require higher dosages of levodopa to correct compared to basic motor circuitry. This could be attributed to differences in dopamine receptor density between cortical and subcortical regions; there are far fewer D_1_ and D_2_ receptors present in the frontal cortex compared to the basal ganglia ^[44,45]^. Still, further exploration, both with subjects receiving higher dosages, and ON-OFF experimental design are required to investigate this speculative hypothesis.

For people presenting with only mild cognitive impairment without any basic motor deficits, uncertainty in motor planning was also observed, although to a slightly lesser degree than the PD cohort. This adds to the suggestion that the planning deficits measured in PD are unlikely to be a consequence of basic motor pathology, which is absent in these MCI patients. The MCI cohort also showed significant prolongation of RT but only minor, insignificant delays of MT compared to healthy controls. This is in agreement with indications from previous studies that the prolonging of RT in this task is not solely a consequence of bradykinesia, but instead inclusive of motor planning deficits or slowness in information processing ^[9]^. In support of this was our finding of a lack of correlation between bradykinesia severity in those with IPD and RT. To separate out contributions to RT prolongation, the lack of correlation between our motor planning measure and RT across all cohorts becomes of particular interest. These results together suggests that, at least in our task, RT is not merely an output measure reflecting the severity of motor planning deficits or bradykinesia, but the prolongation observed is likely also influenced by slowness in other cortical processes, potentially visuospatial or executive circuitry ^[46,47]^.

A key difference to highlight between previous work on simple and choice RT tasks and our study is the freedom of the participant to choose their own grasp, as opposed to the enforcement of an instructed action. Therefore one cannot directly compare RT measurements between such tasks and our study. Of course, the process of motor planning may continue beyond home-pad release and thus our measure of RT alone may not capture the full duration of motor planning. Consequently, the prolongation of MT seen with patient groups in this task likely does not solely reflect the time to execute the reach-to-grasp movement (supported by the lack of correlation between bradykinesia severity and MT) but may incorporate a fraction of planning time as well, perhaps explaining the insignificant but slight delay for MCI subjects ^[48]^. Kinematic comparisons between patient groups of hand and digit posture over the course of the reach flight trajectory would yield a more accurate measurement of planning time and aid elucidation in future work.

Notably, our halfwidth measure did correlate with disease severity for the MCI cohort, with a lower global cognitive score relating to worsened task performance – indicative of a more globally unified pathology. Provided that, as mentioned previously, motor planning deficits are seemingly only observed in individuals with MCI for tasks of higher cognitive load ^[16]^, it could be implied that those who have more severe global impairment might be more sensitive to task difficulty, and despite the physical action of the task being simple, the added ambiguous context and choice freedom might provide sufficient cognitive load to induce minor deficits. The underlying deficits driving motor planning impairment in individuals with MCI could be wide-ranging across the cortex given the range of cognitive deficit profiles included in this study, limiting any definitive mechanistic conclusions for this cohort as a whole. Impairments in task performance could arise from deficits in visuospatial or executive function, however, the uncommon observation of visuospatial deficits in our MCI cohort makes it unlikely that we can solely equate or attribute task performance to this specific behaviour, though it could still be a contributing factor. Similarly, the lack of correlation between executive function and halfwidth further suggests that task performance might not reflect specific functionality in specific brain regions, but perhaps is a reflection of global cognitive load or complex interactions between different functional or disease networks. However, the lack of correlation between this and other MOCA subdomains and task performance could reflect insufficient statistical power given the sample sizes used, rather than an absence of effect.

The pathological neural circuitry underlying the observed deficits in our task could arise differently in PD from MCI. For PD, the wide range of erroneous cortical processes are primarily associated with dysfunctional cortico-striatal-thalamic networks ^[49]^, resulting from progressive loss of dopaminergic neurons in the substantia nigra pars compacta (SNc). By the point of motor symptom onset and thus diagnosis, around half the dopaminergic neurons in the SNc are degenerated ^[50]^, having already resulted in various prodromal non-motor symptoms. This initial degeneration is associated with significant loss of cortico-cortical projecting pyramidal neurons in the pre-supplementary motor area (pre-SMA) ^[51]^, an area vital for the selection of and preparation for movements required, potentially explaining the impairments observed in our study. Furthermore, early disease evolution in the prodromal phase is also associated with alterations to motor cortical excitability and plasticity. Specifically, loss of long-term potentiation and depression, reduced intracortical inhibition, and enhanced plasticity induced by paired associative stimulation ^[52,53]^. In keeping with previous discussion points, the extent of alterations have inconsistent correlations with MDS-UPDRS scoring ^[52]^. These changes could be explained by dopamine deficiency decreasing thalamo-cortical output ^[52]^; such mechanisms might also influence pre-SMA functioning. A dopamine deficiency upstream affecting pre-SMA would align with our earlier discussion of the relationship between levodopa dosage and task performance, with higher dosages needed to influence regions of lower dopamine receptor density. However, it is important to consider that cellular vulnerability, induced by dopamine depletion/dysregulation, glutamate excitotoxicity, protein aggregation, or neuroinflammation, are also credible mechanisms of pre-SMA dysfunction in addition to decreased thalamo-cortical output ^[51,54]^.

One major input needed by pre-SMA and the wider motor planning network in order to achieve successful motor planning is appropriate sensory feedback. For a task whereby grasp action is driven by comfort preferences ^[55,56]^, one might expect that after performing a less comfortable movement, feedback would facilitate adaptive planning over the course of the experiment ^[57,58]^, illustrated by a reduction in uncertainty over time. One contributor to the deficits observed in our task might therefore be attributed to impaired proprioceptive sensory feedback; precedent for the presence of sensory deficits in PD comes predominantly from freezing of gait and reaching studies ^[59-62]^. Furthermore, altered perception of their own movements ^[59,63]^ or of the target object itself ^[64,65]^, would lead to suboptimal motor planning for the task at hand ^[66]^. It remains to be explored whether those individuals who show greater sensory deficits would consequently perform worse at motor planning in ambiguous contexts.

Impaired motor planning may also arise as a consequence of suboptimal internal models of grasping in ambiguous settings caused by cerebellar dysfunction. If the prediction of sensory consequences is impaired, this could lead to greater uncertainty in grasp choice and thus increased variability in task performance. Indeed, there is increasing evidence to suggest that certain areas of the cerebellum are involved in cognitive and motor network dysfunction in both a compensatory and contributory role in PD ^[67-69]^. However, a specific link to the cerebellum’s possible contribution to dysfunctional motor planning in PD remains to be explored.

To help disentangle the different mechanisms outlined here, an additional future direction could be to correlate performance in our task with different decision making tasks that do not require a sensorimotor role but show deficits in PD ^[70]^. This would help yield insight into whether the greater uncertainty in motor planning here arises due to general decision noise, or dysfunctional circuitry specific to sensorimotor ambiguity such as sensory integration and internal modelling.

Our study has several limitations. Firstly, our use of the term ‘motor planning’ has a broad construct, defined from a complex entanglement of its underlying components which we are unable to differentiate between with our methodology, limiting the specificity of the description of deficits observed. Further to this, our measure of motor planning uncertainty could be influenced by general decision noise or indecisiveness, with the need for future studies to disentangle the potential sources of choice ambiguity. Finally, this study was not pre-registered.

Our approach in assessing motor planning deficits in a reach-to-grasp task demonstrated that people with PD show greater uncertainty in ambiguous contexts compared to healthy controls. Furthermore, our finding that disease severity was not an influencing factor revealed the independency of pathological motor planning circuitry from basic motor dysfunction. The robust performance of our measure as a pathology classifier for those in the earlier stages of disease progression showcases the utility of it as a potential diagnostic marker, aiding the formation of a wider clinical picture of the manifestations of Parkinson’s disease.

## Acknowledgements

This work was supported by grants BB/P006027/1 (AK), BB/Y000625/1 (AK), Newcastle Neuroscience Fund (NJM). The funders had no role in study design, data collection or interpretation, nor in the decision to submit the work for publication. The authors thank Mr Norman Charlton for mechanical engineering of the experimental setup, and Prof. Stuart Baker for designing the contact circuit and initial Sequencer scripts.

## Author Contribution

NJM, HK, SC, DSS and AK designed the study. NJM, DSS and AK programmed the behavioural paradigm. NJM collected and processed data, performed analyses, prepared all figures and wrote the manuscript. SB, SS and SM collected data. AK performed analyses. SC, HK, DSS and AK provided resources. DSS and AK edited the manuscript and supervised. All authors read and approved the final manuscript.

## Data Availability

All data generated or analysed during this study are included this article’s supplementary information files.

